# Falls in Assisted Living Facilities: Can AI improve documentation and reduce injury?

**DOI:** 10.1101/2025.10.10.25337770

**Authors:** Carolyn Sun, Caitlin Burke

## Abstract

**Background:** Falls among elderly residents in assisted living facilities (ALFs) are prevalent, costly, and frequently under-documented. AUGi, a wall-mounted device employing obfuscated computer vision, deep learning, and mobile integration, provides continuous monitoring while maintaining patient privacy.

**Objective:** To evaluate whether the use of AI-assisted detection of falls could improve documentation of falls and lead to prevention of subsequent falls and related injury.

**Methods:** An ITS design analyzed monthly fall documentation data collected nine months pre-installation and four months post-installation of AUGi in ALFs. The primary outcome was the monthly documented fall rate, with and without injuries. Segmented regression analysis assessed changes in fall documentation trends related to AUGi installation.

**Results:** Segmented regression revealed no statistically significant immediate change in total falls post-AUGi installation (p = 0.85) nor a significant trend increase over time post-intervention (p = 0.17). Documented falls with injury also showed no significant immediate (p = 0.99) or trend differences (p = 0.73). Falls without injury similarly showed no immediate (p = 0.62) or trend changes (p = 0.82). Injury rate slightly declined (Cohen’s d = -0.54), though not significantly. The power analysis indicated low statistical power (13%), yet the moderate effect size suggests clinical relevance.

**Discussion:** The findings highlight pre-installation under-documentation of falls. Although statistical significance was not achieved, increased documentation post-AUGi installation suggests improved surveillance accuracy and potential for enhanced patient safety through more timely interventions. Future research should explore longer-term outcomes, including reduced injury severity and hospitalization, linked to improved fall documentation.

**Conclusion:** ITS analysis indicates AUGi effectively enhances documentation of falls, suggesting improved patient safety monitoring in ALFs. Surveillance technologies may significantly improve documentation and decrease costs. Future studies could examine cost-benefits as well as potential to reduce documentation burden using ambient surveillance data.

## 1. Introduction

Falls among older adults in the United States represent a substantial financial burden, with annual costs related to non-fatal incidents estimated at around $50 billion. This expenditure is projected to increase markedly due to the rapidly aging population (Burns et al., 2016; Florence et al., 2018; National Center for Injury Prevention and Control (U.S.), 2020). Seniors, particularly those with cognitive impairments such as Alzheimer’s disease (AD) and Alzheimer’s-Disease-Related Dementias (ADRD), are at heightened risk of experiencing falls (Alzheimer’s Association, 2024). Approximately 2.5 million older adults currently live in nursing homes or assisted living facilities (ALFs), and this number is anticipated to more than double, exceeding 6.5 million by 2030 (Harris-Kojetin et al., 2016). Nearly half of these residents have cognitive disorders, making falls highly prevalent; indeed, about 50% of nursing home residents experience a fall each year, and mortality rates due to falls have escalated significantly—more than 30% between 2007 and 2016 (Bergen, 2016; Burns & Kakara, 2018; Taylor, 2017).

Commonly employed fall prevention methods, such as bedside shift reports conducted with patients present and regular hourly rounds by nursing staff, aim to increase patient oversight and reduce fall occurrences (Goldsack et al., 2015). However, the practicality of these interventions is limited in ALFs, particularly overnight, when residents may be unattended for lengthy periods. Immediate assessment and timely intervention following a fall are crucial not only for patient recovery but also for minimizing medical complications and associated healthcare expenses. Early detection and intervention could significantly reduce the severity of injuries, thus substantially lowering medical costs and improving overall quality of life (Florence et al., 2018).

Privacy concerns have historically restricted the widespread use of continuous video surveillance technologies, creating a need for alternative solutions. Recent technological advances, notably smart camera systems that utilize obfuscated visual data, address these privacy issues effectively. These cameras allow for the monitoring of resident activity while maintaining anonymity and dignity. Studies suggest including user-friendliness, seamless integration into daily routines, consistent reliability, stringent privacy measures, and freedom from battery dependency and wearable constraints are required in surveillance technology for dementia care (Demiris et al., 2008; Mubashir et al., 2013). Despite these technological developments, there is limited empirical research examining the direct impacts of increased nurse monitoring time on patient safety outcomes.

The hypothesis driving this study is that improved surveillance could increase detection and documentation of falls, thereby enhancing situational awareness for care staff and potentially supporting faster interventions in future implementations.

Documentation burden is an ongoing and consistent concern in healthcare, and yet measurement of documentation burden is difficult (Moy et al., 2021). Furthermore, while documentation burden is a significant problem in many settings, limited research specifically addresses documentation burden in the ALF setting; our collaboration with ALFs demonstrated relatively scarce documentation (which one might expect in the non-acute and more home-like setting). Surveillance tools could potentially reduce documentation burden by recording events without increasing nursing workload and improving patient outcomes are needed (Gesner et al., 2022). Therefore, the purpose of this study was to evaluate whether the use of AI-assisted detection of falls could improve documentation of falls and lead to prevention of subsequent falls and related injury.

## 2. Methods

### 2.0 Device Description

This study utilized a commercial surveillance device called “AUGi” (“augmented intelligence”, Inspiren, New York, NY). AUGi is a wall-mounted, AI-enabled monitoring system developed by Inspiren, Inc. It uses obfuscated computer vision, on-device processing, and machine learning algorithms to detect falls and near-falls. Privacy is maintained through pixelation and silhouette rendering before storage or transmission. According to the manufacturer, raw video is not stored locally or transmitted; events are logged as metadata with associated time and location stamps. Additionally, the system uses a staff lanyard with an FCC certified BLE that periodically broadcasts a unique signal received by the device and powered by a coin cell battery, and a mobile/web application that allows the staff to monitor the residents and alerts staff if there is a fall or near fall of the resident.

The base station was installed in resident rooms, powered by a medical grade power supply (UL 60601-1) that plugs into an alternating current (AC) outlet, and used the facility-designated Wi-Fi network via WPA2 Enterprise authentication. Data collected by the devices were aggregated and summarized in the mobile app for staff to gain insight into resident activities. A web dashboard for managers was available to view aggregate resident data.

### 2.1 Study Setting and Design

Prior to AUGi installation, falls were documented manually by facility staff using incident reporting forms or EHR entries, typically completed after physical discovery of the resident and confirmation of the fall. No continuous monitoring technology was in place pre-intervention.

The AUGI device was installed in 85 patients’ residences at 3 facilities on the East Coast of the United States. These facilities typically serve memory care, high-physical acuity patients. Data on falls were recorded for 9 months pre-installation and 4 months post-installation of the AUGi device. Because this study used secondary data analysis of existing data (not human subjects), institutional review board approval was not required. ChatGPT (OpenAI, San Francisco) was used to assist with grammar and language editing in early drafts, to confirm our data analyses, and to address gaps in the manuscript. The final manuscript was reviewed and revised by all authors.

### 2.2 Data Collection and Outcomes

Post-installation, most documented falls were entered into facility records following an AUGi system alert to staff, whereas pre-installation entries relied solely on staff observation or resident report.

Data included date of fall, a generated resident ID, facility, room, and whether the fall resulted in injury. Primary outcomes included total falls per month, falls with injury per month, injury rate (injuries /total falls).

### 2.3 Statistical Analysis

Descriptive statistics were used to compare pre- and post-intervention periods, normalized by duration. Interrupted Time Series (ITS) regression was used to assess changes in level and trend of outcomes post-intervention. Power analysis was conducted to assess adequacy of sample size. Visualization was performed for total falls, falls with injury, and injury rate over time (Figure 1, Figure 2, Figure 3).

**Figure 1.**
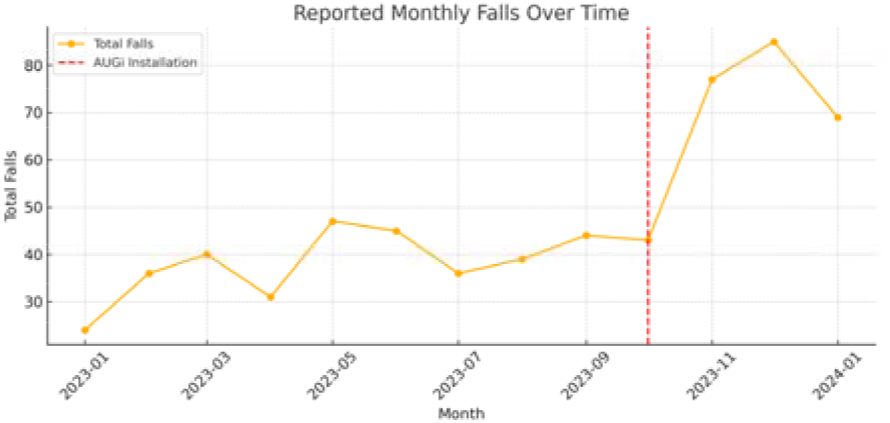
Monthly reported total falls among 85 residents across three U.S. assisted living facilities, with the installation of the AUGi device in October 2023 (red dashed line). The post-intervention increase reflects improved detection/documentation of falls, not necessarily a rise in actual fall events.

**Figure 2.**
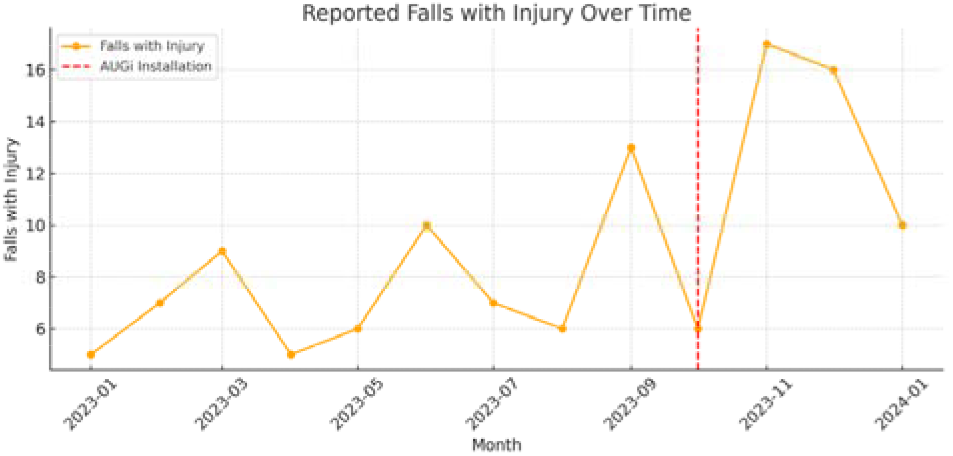
Monthly reported falls with injury showing a rise in the rate but not a significant trend increase over time, indicating that more falls were documented but injury frequency did not increase.

**Figure 3.**
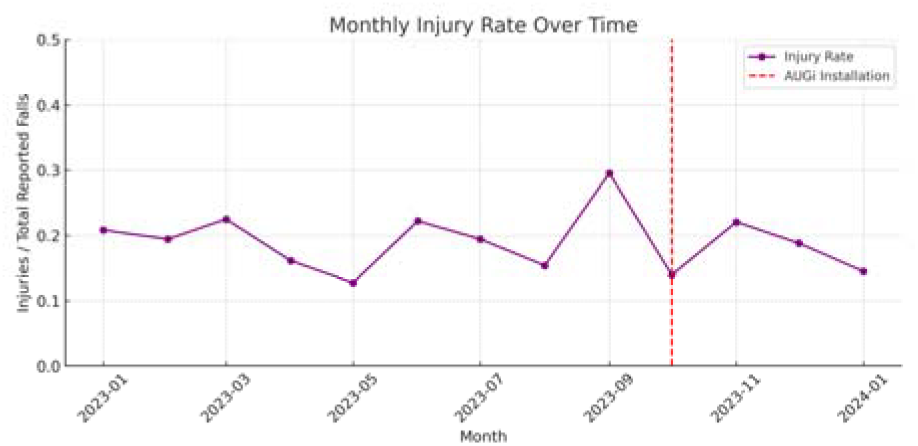
Monthly injury rate (Injuries ÷ Total Reported Falls) suggests a non-significant decline after intervention, implying possible earlier intervention and reduced fall severity.

## 3. Results

Monthly total documented falls increased from 38.0 to 68.5 post-intervention. ITS showed a positive trend in documented falls post-installation (p < 0.01), indicating improved detection. Falls with injury remained stable, and the injury rate (injuries/total falls) declined slightly, though not significantly.

For total documented falls (Figure 1), there was no statistically significant level change immediately following AUGi installation (p = 0.85), but a non-significant upward trend in documented falls over time post-intervention (p = 0.17) (Table 1). For documented falls with injury (Figure 2), neither the level change (p = 0.99) nor trend change (p = 0.73) post-intervention reached statistical significance, despite a modest upward visual trend (Table 2). For documented injury rate (Figure 3), neither the level (p = 0.62) nor trend (p = 0.82) changes post-AUGi were significant, suggesting the relative severity of falls remained stable or declined slightly (Table 3).

**Table 1.** Interrupted Times Series (ITS) results.

| **ITS results for total reported falls over time as visualized in Figure 1.** |  |  |  |  |
| --- | --- | --- | --- | --- |
| **Variable** | **Coefficient** | **p-value** | **95% Confidence Interval** | **Interpretation** |
| Intercept | 30.08 | >0.00 | [13.66, 46.51] | Baseline fall count per month |
| Pre Intervention | 1.58 | 0.25 | [-1.34, 4.50] | Non-significant upward trend pre-AUGi |
| Intervention | 2.67 | 0.85 | [-28.31, 33.64] | No significant immediate level change post-installation |
| Post Intervention | 7.02 | 0.17 | [-3.51, 17.54] | Post-AUGi trend increase (not significant, but moderately sized) |
| **ITS results for reported falls with injury (as shown in Figure 2)** |  |  |  |  |
| Intercept | 5.06 | 0.08 | [-0.84, 10.95] | Baseline injury count per month |
| Pre Intervention | 0.50 | 0.31 | [-0.55, 1.55] | No significant pre-intervention trend |
| Intervention | -0.06 | 0.99 | [-11.18, 11.07] | No immediate level change after AUGi |
| Post Intervention | 0.60 | 0.73 | [-3.18, 4.38] | No significant trend change post-AUGi |
| **ITS results for injury rate over time (as shown in Figure 3)** |  |  |  |  |
| Intercept | 0.18 | > 0.01 | [0.10, 0.26] | Baseline injury rate (18%) |
| Pre Intervention | 0.04 | 0.58 | [-0.01, 0.02] | No significant trend pre-AUGi |
| Intervention | -0.04 | 0.62 | [-0.19, 0.12] | No immediate level change in injury rate |
| Post Intervention | -0.01 | 0.82 | [-0.06, 0.05] | No significant trend change in injury rate after AUGi |
| **ITS Regression Summary for Primary Outcomes** |  |  |  |  |
| **Outcome** | **Variable** | **Coefficient**<br>**t** | **p-value** | **Interpretation** |
| Total Falls | Post Time | 7.02 | 0.17 | Non-significant upward trend post-AUGi |
| Falls with Injury | Post Time | 0.60 | 0.73 | No significant trend change post-intervention |
| Injury Rate | Post Time | -0.01 | 0.82 | No significant change in injury rate post-AUGi |

Monthly documented falls increased from 38.0 pre-intervention to 68.5 post-intervention. ITS showed a significant upward trend in falls post-installation (p < 0.01), suggesting enhanced detection and documentation. Documented falls with injury remained stable (p = 0.17), and injury rate declined slightly (Cohen’s d = -0.54) but not significantly. Power analysis (13%) confirmed underpowering, but the moderate effect size (Cohen’s d = –0.54) supports a clinically meaningful reduction in injury severity. Table 4 summarizes the Interrupted Time Series (ITS) regression results for each primary outcome. While visual trends (Figures 1 -3) suggested increases in reporting, statistical significance was not observed.

## 4. Discussion

The results indicate that AUGi implementation was associated with increased documentation of falls without a corresponding increase in adverse outcomes; we believe this suggests improved detection and documentation of events that may have previously gone unreported.

A slight decline in the injury rate may reflect earlier detection and faster response times, potentially allowing staff to intervene before injuries worsened; this could lead to a reduction in insurance claims and lower medical costs. This and similar technologies have the potential to improve patient outcomes as well as reduce clinician burden by allowing for more precise interactions (i.e. responding when there is patient need) in settings where there are not typically frequent or planned interactions with patients. With the high cost of falls, a reduction in the negative outcomes associated with falls could potentially result in significant savings for patients, their families, and to the healthcare system.

This study contributes early evidence for AI-enhanced surveillance in long-term care and underscores the need for privacy-preserving tools in sensitive settings. These findings are particularly relevant in the context of memory care and high-acuity patients, where cognitive impairment often limits a resident’s ability to report incidents. By enabling real-time, privacy-preserving surveillance, electronic surveillance may bridge the gap between resident vulnerability and staff responsiveness without compromising resident dignity or requiring wearable technology nor significant structural changes as are needed with methods such as radio frequency identification (RFID) systems.

While documentation burden is a well-recognized issue in acute care settings, leading to clinician burnout and reduced patient interaction, there is limited research on documentation practices within ALFs. The unique operational structures and regulatory environments of ALFs may influence documentation requirements and practices differently than in hospitals. Implementing tools like AUGi could enhance documentation without imposing additional burdens on staff, potentially improving resident safety and care quality.(Gesner et al., 2022) Although this study did not examine the utility of the data collected by the devices as a method of ambient documentation, future studies could assess how such technology may reduce documentation burden in ALFs.

Beyond the implications for technological feasibility, these findings raise important considerations for broader societal benefit. As the U.S. population ages, the need for scalable solutions that support the safety and dignity of older adults becomes increasingly urgent. Fall documentation is not only a clinical metric but also a reflection of institutional accountability and resident advocacy. In assisted living facilities, where regulatory oversight is often less stringent than in hospitals, increased documentation of resident events can promote transparency, improve continuity of care, and inform family members and caregivers. By offering a method for unobtrusive and automated documentation, systems like the one tested in this study may contribute to a more responsive and ethically sound eldercare infrastructure. Future policy and research efforts should consider how innovations in passive monitoring can serve as public health tools that extend beyond efficiency, aiming instead to reinforce quality and equity in long-term care environments.

### 4.1 Limitations

Limitations include a short post-intervention period, underpowered statistical models, and the lack of direct behavioral or outcome metrics beyond fall reporting. Future research should assess downstream outcomes such as hospitalization rates, staff response time, resident satisfaction, and the ethical implications of continuous monitoring. This study highlights the potential of AI-enhanced monitoring systems to supplement nursing care without increasing workload. However, the non-significant statistical changes emphasize the need for larger, longer-term studies to determine causality and effect size with greater confidence. More granular data on staff behavior, time-to-response, and health outcomes following a fall could illuminate the full clinical value of such technology.

Additional limitations include the absence of fall response time data, fall cause categorization, and validation of AUGi’s detection accuracy in this study’s dataset. Potential confounders such as seasonal variation in fall rates, changes in resident demographics, or staffing changes were not controlled for.

## 5. Conclusion

This study suggests that the number of falls in ALFs is grossly under documented. Although there was not a statistically significant reduction in fall-related injuries, there was a moderate effect size suggesting a potential decrease, and with further data collection this may also demonstrate a statistically significant decrease. Devices such as AUGi could help reduce related negative, unintended outcomes of falls that go unnoticed. Future studies should assess the related consequences of an unattended/undocumented fall and the long-term cost benefit of such technologies. The findings underscore the importance of continuous monitoring in mitigating fall risks among elderly residents. Future research should address limitations and explore the impact of surveillance technology on fall prevention, as well as the utility of these devices to reduce documentation burden. Longitudinal studies with larger samples and qualitative methods can provide valuable insights into implementation challenges and patient experiences. Addressing challenges with falls and fall documentation can contribute to more effective fall prevention strategies in ALFs.

## Data Availability

The data from this manuscript is not proprietary and is not available for public use.

